# Documented Pain Relief After Emergency Department Headache Treatment Is Not a Stable Outcome: Reassessment Timing, Missingness, and Score Selection

**DOI:** 10.64898/2026.07.05.26357324

**Authors:** Alon Gorenshtein, Yosef Adiniaev, Tom Liba, Eyal Klang, Oved Daniel

## Abstract

**Background:** Whether a patient’s pain improved after emergency department (ED) treatment is read from the record to benchmark EDs, compare drugs, and label research outcomes. It is interpretable only if a post-treatment score is recorded, appropriately timed, and chosen by a fixed rule; its stability across these choices is unknown.

**Methods:** Retrospective measurement study of adult headache visits in a de-identified ED database (MIMIC-IV-ED, 2011-2019). Among treated visits, we quantified reassessment completeness by time window, estimated meaningful relief (a reduction of at least 2 points) under score-selection rules and missing-data assumptions, tested whether reassessment was predictable at treatment, and compared headache with other painful presentations.

**Results:** Among 19,501 visits (15,273 patients), 13,682 (70.2%) were treated. A post-treatment pain score appeared at any time for 77.1% (95% CI, 76.4 to 77.8), but within 2 hours of the analgesic for only 47.9% and within 1 hour for 27.5%. Meaningful relief was 66.9% using the first post-treatment score but 81.0% and 83.4% using the last or lowest score; it was 67.5% under inverse-probability weighting and could be bounded only between 51.8% and 74.4%. Whether a score was recorded was weakly predictable at treatment (area under the curve, 0.566) and unrelated to baseline pain. Completeness was similar across headache strata and comparator painful presentations. In an independent ED (MC-MED, a different EHR), the score-selection effect replicated: relief rose from 71.1% (first) to 80.6% (last) and 83.4% (lowest).

**Conclusions:** Documented pain relief after ED headache treatment was not a stable outcome: it varied with the reassessment window and score-selection rule, was not point-identified for unreassessed patients, and behaved like other painful ED presentations. Programs and research that use documented relief should prespecify the reassessment window, score-selection rule, completeness denominator, and a missing-data range, and favor protocol-timed reassessment.

## Introduction

Treating acute pain is one of the most common tasks in the emergency department (ED), and whether the patient’s pain improved is treated as a core measure of whether the visit succeeded.[1,2] That question is increasingly answered from the record rather than the bedside: documented pain change after analgesia is used to benchmark EDs against one another, to compare analgesic agents, and to label outcomes for emergency-care research and operational dashboards, often without the time anchoring and protocolized reassessment of a trial.[2,3,4] In the ED, however, that documentation is produced under crowding and by nurse-driven reassessment rather than on a fixed schedule.[3,6] Headache is the index presentation for this problem because relief is the core response endpoint there, measured at fixed times in ED migraine trials and society treatment assessments and used as the comparator in the case against routine opioids, so headache-response labels read from the record are the ones most likely to be over-read.[12,13]

A documented relief measure is interpretable only under conditions that a trial enforces but routine ED care does not. The post-treatment pain score must be recorded, it must be recorded at a clinically interpretable time, and when several scores exist a rule must decide which one counts. Each of these is a documentation and analysis choice rather than a property of the patient or the drug. Prior work has shown that pain reassessment after analgesia is documented in only a minority of ED encounters and that the numeric pain scale is a coarse, heaped instrument, but it has treated these as quality or instrument problems rather than as threats to the validity of a relief measure read from the record.[3,5,6,7,8] Whether the measure itself is stable across clinically plausible definitions, applied to the same treated patients, has not been examined; it cannot be examined with the single triage pain value in national survey data, but is measurable in detailed ED records with serial scores and timestamped dispensing.[9,10]

We used a large de-identified ED database to test the stability of documented pain relief after ED headache treatment. We quantified reassessment completeness across clinically relevant time windows, measured how far the relief estimate moved with the post-treatment score-selection rule and with assumptions about unmeasured patients, asked whether reassessment was predictable at the time of treatment, and tested whether the pattern was specific to headache or general to ED pain by comparing it with back, abdominal, and renal-colic presentations. We hypothesized that documented relief would be unstable across these clinically plausible definitions even when applied to the identical set of treated headache visits.

## Methods

### Study Design and Setting

This was a retrospective measurement study of adult ED visits for headache in the MIMIC-IV-ED module (version 2.2), a de-identified database of visits to one US academic ED (Beth Israel Deaconess Medical Center) between 2011 and 2019. Reporting followed STROBE with the RECORD extension; the reassessment-prediction component followed TRIPOD (eMethods; item map in eTable 9). The quantity of interest was a property of the medical record, namely whether, when, and how the response to analgesia was documented. No causal effect of treatment on a clinical outcome was estimated.

### Cohort and Comparators

Headache visits were identified by a triage chief-complaint term (headache, head ache, migraine, cephalgia, or cephalalgia) or an ED diagnosis coded as a primary or nonspecific headache disorder (ICD-9 346.x, 307.81, 339.x, or 784.0; ICD-10 G43, G44, or R51; eTable 1). Analyses were restricted to adults. The treated subcohort received at least one qualifying analgesic or abortive agent (opioid, nonsteroidal anti-inflammatory drug [NSAID] or ketorolac, acetaminophen, dopamine-antagonist antiemetic, triptan, dihydroergotamine, or intravenous magnesium; eTable 2). Prespecified strata were migraine-coded versus nonspecific headache, chief-complaint versus diagnosis identification, discharged-home visits, and a headache-directed-therapy subcohort (dopamine antagonist, triptan, dihydroergotamine, NSAID or ketorolac, or magnesium). To judge whether documentation patterns were specific to headache, comparator cohorts of other common painful presentations (back, abdominal, chest, and renal-colic pain) were constructed by the same chief-complaint method.

### The Documented Response Endpoint

The index treatment time was the earliest qualifying dispense; because the dispensing table records cabinet-removal rather than administration time, it was treated as a documented care event. The baseline pain score was the last numeric 0 to 10 value at or before the index time. A documented reassessment was a numeric pain value after the index time within the same ED stay; because anytime documentation is the most permissive definition, completeness was also computed within 30, 60, 90, 120, and 180 minutes. The documented response was baseline minus a post-index value, with meaningful relief at a reduction of at least 2 points (primary) or at least 30%. Because more than one post-index score could exist, the relief rate was computed under three score-selection rules, using the first, the last, and the lowest (best) post-index value within the same ED stay; the last and lowest rules were stress tests of endpoint stability rather than proposed endpoints. Response analyses required an interpretable baseline of 1 to 10.

### Statistical Analysis

Completeness was a proportion with a 95% Wilson confidence interval (CI). The population relief rate was estimated by complete-case analysis, by inverse-probability-of-reassessment weighting (IPW) under a missing-at-random (MAR) assumption (weights truncated at the 99th percentile; diagnostics in eTable 7), and by missing-not-at-random (MNAR) bounding sweeping the assumed relief among never-reassessed patients across its full range, with model-based imputation under MAR and fixed MNAR offsets. The presence of a reassessment was modeled by logistic regression in two specifications: an at-treatment model using only factors known at or before the index dispense, and a full model adding the later visit course (ED length of stay, total medication classes, any opioid or parenteral agent); discrimination used 5-fold cross-validated out-of-fold AUC with a 2,000-resample bootstrap CI and the Brier score, with a gradient-boosted comparison by paired bootstrap, and adjusted odds ratios with Benjamini-Hochberg correction (eTable 3; eFigure 2). Because patients could contribute multiple visits, key estimates were repeated using one visit per patient and a patient-clustered bootstrap. Categorical and continuous group differences used the chi-square test with Cramer V and the Kruskal-Wallis test with epsilon-squared. The threshold was *P* < .05 (two-sided). Analyses used Python 3.9 (pandas 2.3, scikit-learn 1.6, SciPy 1.13, statsmodels 0.14); seeds were fixed.

### External Validation

To test whether the core measurement findings depended on a single center or EHR, the three primary analyses (reassessment completeness, the score-selection relief estimate, and the at-treatment predictability of whether a reassessment occurs) were repeated unchanged in an independent dataset, MC-MED (PhysioNet, version 1.0.0), a de-identified ED database from a separate US academic medical center spanning approximately 2020 to 2022 on a different EHR.[14] Headache visits were identified by triage chief complaint and the treated subcohort received a qualifying analgesic, matching the primary cohort definitions; serial pain scores were drawn from the MC-MED numerics stream. The index treatment time in MC-MED was the medication administration time (the First_admin_time field) rather than the dispense time used in MIMIC-IV-ED, and MC-MED date-shifting preserves within-visit intervals, so treatment-to-reassessment timing was valid. The same estimators, score-selection rules, and cross-validated discrimination procedures were applied.

### Ethics

MIMIC-IV-ED was approved by the institutional review boards of the Beth Israel Deaconess Medical Center and the Massachusetts Institute of Technology, with a waiver of individual consent because records are de-identified. Access was through credentialed PhysioNet authorization. This secondary analysis did not require additional review board approval.

## Results

### Cohort

A total of 19,501 adult ED visits for headache by 15,273 patients were identified (median age, 42 years; 69.8% female; Table 1); 2,407 patients (15.8%) contributed more than one visit. Triage acuity was concentrated at Emergency Severity Index (ESI) levels 2 and 3, the median triage pain score was 7 of 10, and most visits were discharged home (74.8%). A qualifying analgesic or abortive agent was dispensed in 13,682 visits (70.2%; 95% CI, 69.5 to 70.8), which formed the treated subcohort.

**Table 1.** Characteristics of the emergency department headache cohort.

| Characteristic | Value (N = 19,501) |
| --- | --- |
| Patients, No. | 15,273 |
| Age, median (IQR), y | 42 (28-56) |
| Female, No. (%) | 13,612 (69.8) |
| Race and ethnicity, No. (%) |  |
| White | 9,594 (49.2) |
| Black | 5,207 (26.7) |
| Hispanic | 2,301 (11.8) |
| Asian | 819 (4.2) |
| Unknown or other | 1,580 (8.1) |
| Ambulance arrival, No. (%) | 4,856 (24.9) |
| Triage acuity (ESI), No. (%) |  |
| 1 (most acute) | 878 (4.5) |
| 2 | 6,057 (31.1) |
| 3 | 11,984 (61.5) |
| 4-5 (least acute) | 528 (2.7) |
| Triage pain score, median (of 10) | 7 |
| Disposition, No. (%) |  |
| Discharged home | 14,587 (74.8) |
| Admitted | 3,842 (19.7) |
| Left without being seen | 449 (2.3) |
| ED length of stay, median (IQR), h | 5.6 (3.9-8.0) |
| Received analgesic or abortive (treated), No. (%) | 13,682 (70.2) |

Abbreviations: ED, emergency department; ESI, Emergency Severity Index; IQR, interquartile range. Acuity was missing in 54 visits (0.3%).

### Whether the Endpoint Exists Depends on the Time Window

A post-treatment pain score was eventually recorded for most treated visits, but a clinically timed reassessment was the exception. Of 13,682 treated visits, 10,553 (77.1%; 95% CI, 76.4 to 77.8) had a post-treatment score at some point in the stay, an any-time definition that is the most permissive available and should not be read as reassuring. Within clinically meaningful windows, documented reassessment was 60.7% within 180 minutes, 47.9% within 120 minutes, 38.5% within 90 minutes, and 27.5% within 60 minutes of the analgesic (Figure 1A); the median time to first reassessment was 91 minutes (IQR, 37 to 164). Completeness was stable across migraine-coded, nonspecific, headache-directed-therapy, and discharged-home strata (any-time 76 to 79%; 2-hour near 48% throughout; eTable 4), and unchanged in patient-clustered and one-visit-per-patient analyses.

**Figure 1.**
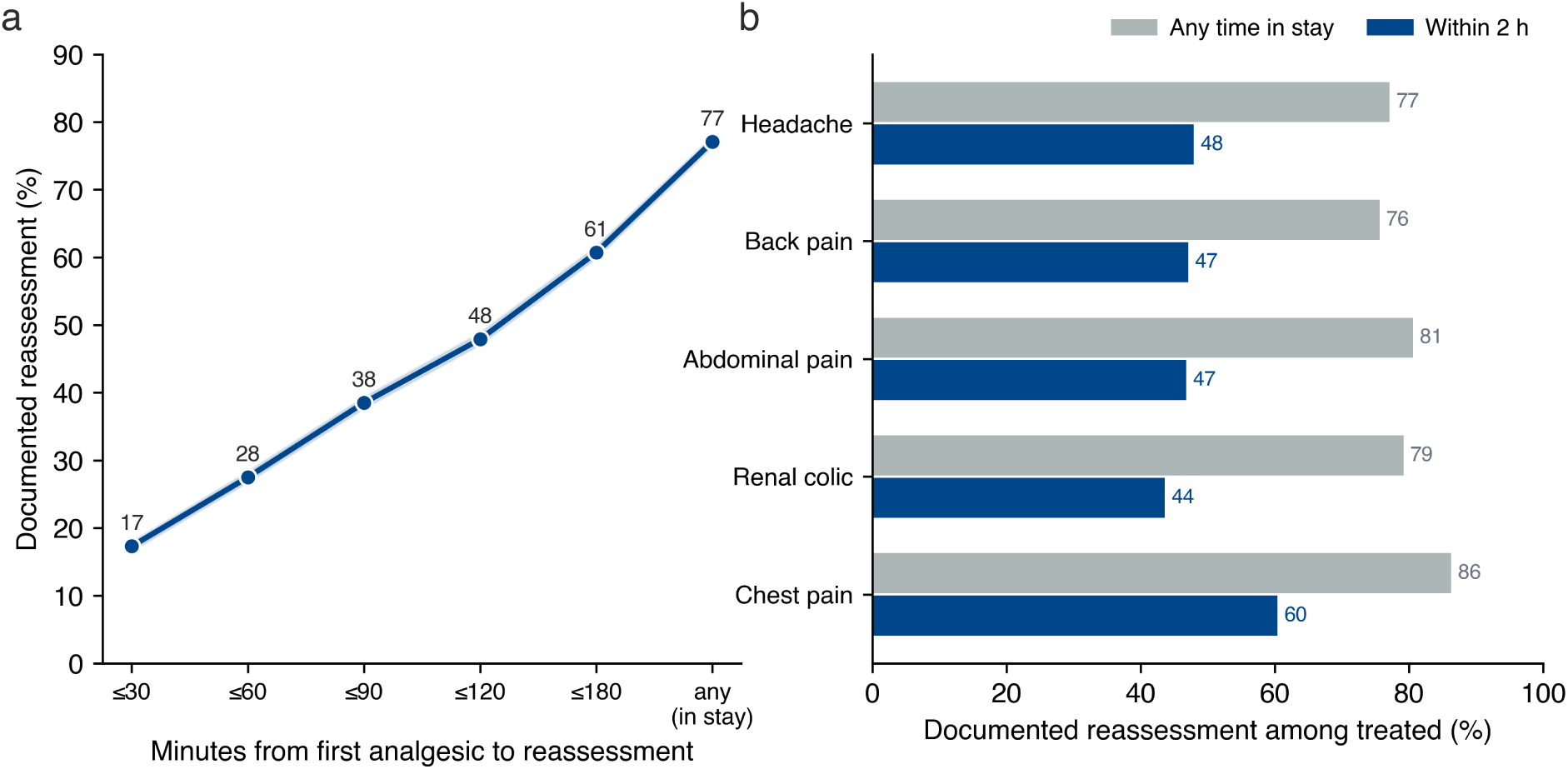
Timing and cross-presentation completeness of post-treatment pain reassessment. (A) Among treated headache visits, the percentage with a documented post-treatment pain score within increasing time windows after the first analgesic and at any time before disposition; the shaded band is the 95% Wilson confidence interval. (B) Any-time and within-2-hour reassessment among treated visits for headache and for comparator painful presentations constructed identically.

### The Relief Estimate Shifts Across Score-Selection Rules and Missing-Data Assumptions

**Applied to the identical treated cohort, the documented relief rate ranged from about half to more than four-fifths depending only on analysis choices.** Among 11,987 treated visits with an interpretable baseline pain, 9,281 (77.4%) were reassessed and 2,706 (22.6%) had no observable response; excluded visits had a similar reassessment rate (eTable 8). Using the first post-treatment score, 66.9% of reassessed patients achieved a reduction of at least 2 points (mean reduction, 3.4 points); using the last or the lowest documented score instead raised this to 81.0% and 83.4% (Figure 2). This sensitivity was an artifact of repeated measurement. Among reassessed visits the median number of post-treatment scores was 1 (IQR, 1 to 2) and 53.3% had only one, for whom the first and best scores coincide; where multiple scores existed, the best-score rule inflated relief by 28.2 percentage points with two scores and 43.9 with three or more, while first-score relief fell as scores accrued (from 79.1% with one score to 46.7% with three or more), consistent with further measurement being prompted by slower improvement and then capturing a transient low. The last documented score fell a median of 176 minutes (IQR, 95 to 321) after treatment, often near disposition, so neither the last nor the best score is a superior endpoint (eTable 10). The choice of missing-data treatment moved the population estimate less but in interpretable directions: complete-case and IPW estimates were nearly identical (66.9% and 67.5%; weights well behaved, effective sample size 9,162 of 9,281, calibration slope 0.96, all standardized mean differences below 0.01 after weighting; eTable 7), so under MAR the documented figure was approximately unbiased, whereas model-based imputation that lowered relief among the unmeasured by 20 and 40 percentage points returned 63.0% and 58.5%. Allowing the unmeasured relief rate to span its full range, the population proportion with a reduction of at least 2 points was identified only between 51.8% and 74.4%, a band of 22.6 percentage points equal to the unmeasured fraction (eFigure 3). The wide identification bound applied within the migraine, nonspecific, and headache-directed-therapy strata, although the complete-case estimate was higher in migraine-coded visits (74.1%) than in nonspecific headache (66.1%; eTable 6).

**Figure 2.**
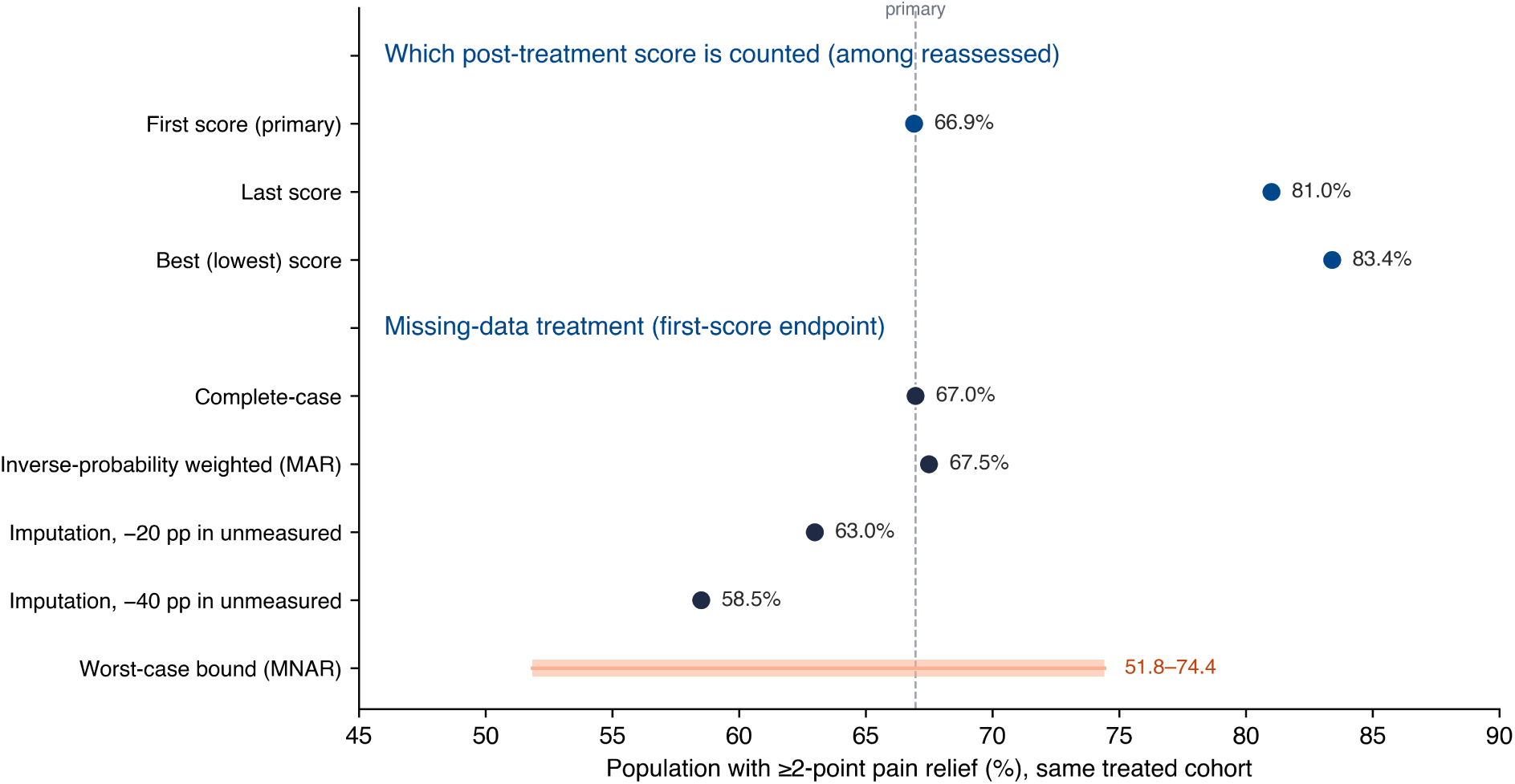
The documented relief endpoint shifts across analysis choices in the same treated cohort. Each point is the population proportion achieving a reduction of at least 2 points under a post-treatment score-selection rule (first, last, or lowest score, among reassessed visits) or a missing-data treatment of the first-score endpoint (complete-case, inverse-probability weighted, or model-based imputation with missing-not-at-random offsets); the salmon bar is the worst-case missing-not-at-random identification bound. The dashed line marks the primary first-score complete-case estimate.

**Figure 3.**
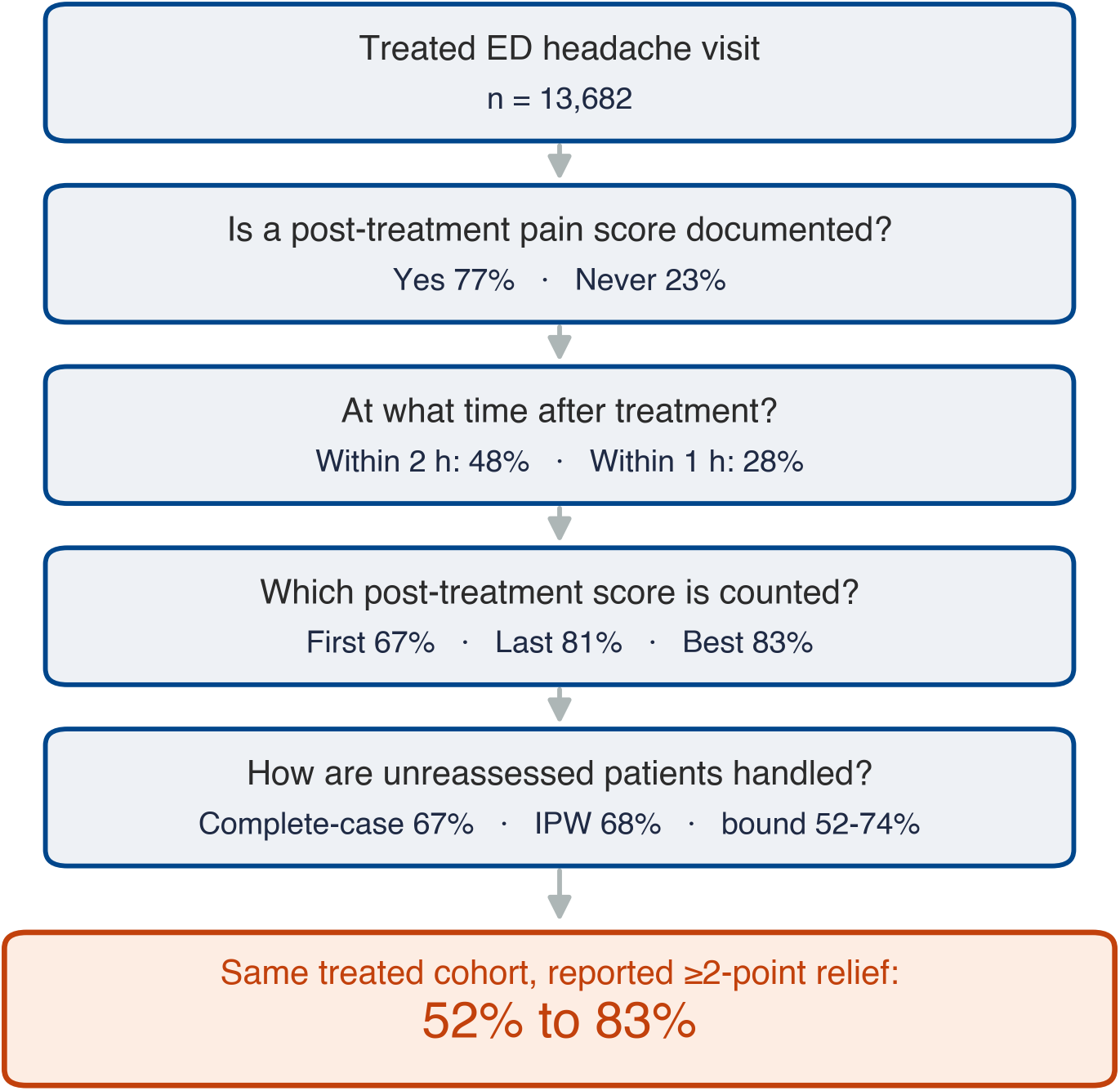
How the documented relief endpoint is constructed from a treated headache visit. Schematic of the documentation and analysis choices that intervene between a treated visit and a reported relief rate: whether a post-treatment score is recorded, at what time, which of several scores is counted, and how unreassessed patients are handled. The numbers at each step are the values observed in this cohort; the same treated cohort yields a reported meaningful-relief rate of 52% to 83% across these choices.

### Whether a Score Is Recorded Is Only Weakly Predictable at Treatment and Unrelated to Pain

Using only information available when the analgesic was given, whether a reassessment would be documented was barely better than chance (at-treatment AUC, 0.566; 95% CI, 0.555 to 0.578); adding the later visit course raised this to 0.636 (95% CI, 0.626 to 0.647), indicating that most of the limited predictability reflected time and activity accrued after treatment rather than selection on the patient. The odds of a documented reassessment increased with ED length of stay (odds ratio [OR], 1.11 per hour; 95% CI, 1.10 to 1.13) and the number of medications (OR, 1.29; 95% CI, 1.21 to 1.38) and was higher on night and evening shifts, but showed no association with baseline pain (OR, 1.01 per point; 95% CI, 1.00 to 1.03; *P* = .26) and no difference by race or ethnicity after adjustment (eTable 3; eFigure 2). A gradient-boosted model did not improve discrimination (difference in AUC, -0.006; 95% CI, -0.011 to -0.002).

### The Pattern Is Not Specific to Headache

In comparator cohorts constructed identically, any-time reassessment was 75.6% for back pain, 80.6% for abdominal pain, 79.2% for renal colic, and 86.3% for chest pain, with 2-hour reassessment of 47.1%, 46.8%, 43.6%, and 60.4% (Figure 1B; eTable 5). Headache (77.1% and 47.9%) sat among the common painful presentations and below the protocol-monitored chest-pain group, indicating that incomplete and time-dependent documentation is a general property of ED pain records rather than a headache-specific deficiency.

### External Validation in MC-MED

The signature score-selection finding replicated in MC-MED. Among 4,843 chief-complaint headache visits, 2,257 were treated with a qualifying analgesic, and in the 211 reassessed responders the documented meaningful-relief rate rose from 71.1% using the first post-treatment score to 80.6% using the last and 83.4% using the lowest (best) score. The first-to-last-to-best ordering and its magnitude matched the primary cohort (66.9%, 81.0%, and 83.4% in MIMIC-IV-ED), and the best-score relief was identical at 83.4%, so the endogenous inflation of relief by the score-selection rule was reproduced in an independent ED on a different EHR. Whether a reassessment occurred was likewise essentially unpredictable from features known at treatment, with a cross-validated AUROC of 0.501 in MC-MED, near the 0.566 observed in MIMIC-IV-ED and close to chance in both. Reassessment completeness was also poor and timing-dependent in MC-MED, although its absolute magnitude did not match MIMIC-IV-ED: a post-treatment pain score appeared at any time before disposition for 11.8% of treated visits (95% CI, 10.6 to 13.2), within 120 minutes for 8.8%, and within 60 minutes for 6.0%, against 77.1%, 47.9%, and 27.5% in MIMIC-IV-ED. The direction was therefore preserved and, if anything, starker, while the completeness rate itself did not port across systems, consistent with the measurability of the endpoint being a function of documentation structure rather than a fixed clinical quantity. Two of the three core findings replicated, including the signature score-selection result, whereas the completeness magnitude was EHR-structure-dependent.

## Discussion

In a large ED headache cohort, documented pain relief read from the record was not a stable outcome. In the identical set of treated visits, the meaningful-relief rate was 67% if the first post-treatment score was counted but 81% to 83% if the last or lowest was counted; a clinically timed reassessment existed within two hours for fewer than half of treated patients; and the population rate could be bounded only across a 23-point range once the never-reassessed were considered. The measure therefore depends on three documentation and analysis choices, each of which a randomized trial fixes by protocol and routine ED care does not: whether a score is recorded, when it is recorded, and which of several scores is selected (Figure 3). Read from the chart, ”did the patient’s pain improve?” reflects these choices more than the drug.

These choices should be kept separate, because they fail in different ways. Reassessment is incomplete, and steeply so at clinically relevant times. Its presence is only weakly predictable at the moment of treatment and is unrelated to how much pain the patient reported; the discrimination that exists comes from the later visit course, consistent with documentation being a byproduct of how long and how actively a visit unfolds rather than a measurement timed to the drug. Given that incompleteness, the population relief rate is not point-identified without assumptions about the unmeasured, although the near-identity of complete-case and inverse-probability-weighted estimates shows that under a missing-at-random assumption the observed adjustment is small. Only this last issue concerns bias, and the larger practical problem is not a proven large bias but the ambiguity of the endpoint itself: the same record yields materially different answers under defensible definitions.

This reframes two strands of prior work. Pain reassessment after analgesia has been documented in a minority of ED encounters and treated as a quality-improvement target, and the numeric pain scale has been described as a coarse, heaped instrument (eFigure 1).[3,5,6,7,8] The present results connect these to the validity of a derived endpoint: when reassessment is time-selected and multiple scores may exist, any single relief figure reflects the time window and the scoring rule as much as the drug. The comparator analysis shows this is not headache-specific neglect; completeness for headache resembled back, abdominal, and renal-colic pain and fell below protocol-monitored chest pain, so the phenomenon is a general feature of ED pain documentation. Headache is nonetheless the consequential demonstration case, because pain relief is the primary response endpoint in ED migraine trials and society treatment assessments and is measured there at anchored times, so headache-response labels drawn from the record without that anchoring are precisely the ones most likely to be over-read.[12,13] The point is distinct from the clinical caution that a favorable analgesic response does not establish a benign headache etiology.[11]

The implications are concrete for emergency care. First, ED pain quality programs and inter-ED or between-drug benchmarking that report the share of treated patients whose pain improved should prespecify and report the reassessment time window, the score-selection rule, the completeness denominator, and a missing-data sensitivity range, because the same data support 67% or 83% relief under choices that are each defensible; an apparent gap in performance between two EDs or two analgesics could reflect a difference in when and how pain was re-scored rather than a difference in care. Second, emergency-care research and learning-health-system models that use a documented pain change as an outcome label inherit this instability, partly learning who was re-measured and which score was kept, so they should model the missingness and fix the scoring rule rather than leave them implicit. Third, the operational reality is that a first-available score recorded a median of 91 minutes after treatment is a weak surrogate for the patient’s response to the drug, which argues for prospective, protocol-timed reassessment when relief is the endpoint rather than reliance on whatever the chart happens to contain.[12]

### Limitations

This study has several limitations. First, the primary analyses draw on a single academic ED, so absolute rates may not generalize; we therefore repeated the three core analyses in an independent academic ED on a different EHR (MC-MED), where the signature score-selection finding replicated closely (first-to-last-to-best meaningful relief of 71.1%, 80.6%, and 83.4%, with the best-score rate identical to the primary cohort) and reassessment remained essentially unpredictable at treatment (AUROC, 0.501). Reassessment completeness was again poor and timing-dependent in MC-MED but at a much lower absolute level (11.8% at any time versus 77.1%), so the completeness rate itself did not port across systems; this non-portability is itself consistent with the central thesis that the measurability of the endpoint reflects documentation structure rather than a fixed clinical quantity. The comparator analysis and the focus on a measurement mechanism that any EHR shares further mitigate the single-center concern. Second, dispensing time approximates administration time, which most affects the tightest windows. Third, headache was identified from chief complaint and ICD codes, which mix primary and nonspecific headache and cannot adjudicate etiology; results were stable across strata, but the migraine-coded subgroup was modest (1,405 treated visits), so the findings apply to EHR-derived headache-response endpoints broadly, with supportive rather than definitive migraine-specific analyses. Fourth, reassessment was a documented numeric value, so verbal or undocumented reassessment is not captured; this is the relevant exposure for record-derived metrics but not for bedside care, and a documented improvement remains valid patient-reported evidence for the individual, distinct from durable relief, recurrence, rescue-medication need, or return visits. Fifth, the best-score rule is not proposed as a superior endpoint; it illustrates how an apparently reasonable extraction choice can mechanically inflate the relief rate, since patients with more scores have more opportunities to record a low value. Finally, the response among the never-reassessed is unobservable, which is the premise of the bounding analysis rather than a remediable gap.

## Conclusions

Documented pain relief after ED headache treatment was not a stable outcome: it varied substantially with the reassessment window and the post-treatment score-selection rule and was not point-identified for unreassessed patients, while behaving similarly to other painful ED presentations. Read from the chart, this measure reflects when and whether a nurse re-scored the pain and which post-treatment score was selected more than it reflects the analgesic. ED pain quality programs, inter-ED and between-drug benchmarking, and emergency-care research that use documented relief should prespecify the time window, the score-selection rule, the completeness denominator, and a missing-data sensitivity range, and should prefer prospective protocol-timed reassessment where relief is the endpoint.

## Supporting information

full appendix

## Data Availability

All data produced are available online at https://github.com/Alon-Gorenshtein/study_headache_unmeasured_relief

