## Supplementary material for "Documented Pain Relief After Emergency Department Headache Treatment Is Not a Stable Outcome: Reassessment Timing, Missingness, and Score Selection": full appendix

This Supplementary Information accompanies the main manuscript. It contains the case-identification and medication value sets, the full multivariable model, reassessment completeness by time window and by clinical stratum, comparator-cohort estimates, the documented relief rate under every missingness assumption and scoring rule, inverse-probability-weighting diagnostics, reassessed-versus-not differences with missing-baseline accounting, and a reporting-guideline item map. It also states the software, code, and data-availability terms.

**Contents**


- S1 Supplementary Methods
- S2 Supplementary Tables (eTable 1 to eTable 10)
- S3 Supplementary Figures (eFigures 1-3)
- S4 Endpoint Codebook
- S5 Code and Data Availability


### S1 Supplementary Methods

#### S1.1 Case identification and comparators

Headache visits were identified by a triage chief-complaint term (regular expression \b(headache|head ache|migraine|cephalgia|cephalalgia)\b) or a primary or nonspecific headache diagnosis (eTable 1); the two criteria overlapped in 9,337 visits. Comparator cohorts (back, abdominal, chest, and renal-colic pain) were built by the same chief-complaint method to judge whether documentation patterns were specific to headache.

#### S1.2 Treatment, reassessment, and time windows

The treated subcohort received at least one qualifying analgesic or abortive dispense (eTable 2). The index time was the earliest qualifying dispense. Because the dispensing table records cabinet-removal rather than administration time, the index time was treated as a documented care event. A documented reassessment was a numeric pain value recorded after the index time within the same ED stay. Because any-time-in-stay documentation is the most permissive definition, completeness was also computed within 30, 60, 90, 120, and 180 minutes of the index dispense (eTable 4).

#### S1.3 Selection models (at-treatment versus full)

The presence of a reassessment was modeled by logistic regression in two specifications. The at-treatment model used only factors known at or before the index dispense: age, sex, race and ethnicity, arrival mode, baseline pain, triage acuity, arrival shift, ED census at the index time, and first-agent class. The full model added factors that accrue over the later visit and are mechanically related to documentation opportunity: ED length of stay, total number of analgesic classes, and receipt of any opioid or any parenteral agent. Comparing the two isolates how much apparent predictability reflects baseline selection versus time at risk. Discrimination used 5-fold stratified cross-validation with out-of-fold predictions, AUROC with a 2,000-resample bootstrap 95% confidence interval (CI), and the Brier score; a gradient-boosted classifier was compared with the standardized-feature logistic pipeline by a paired bootstrap of out-of-fold predictions. Adjusted odds ratios (full model) carried Benjamini-Hochberg-adjusted P values (eTable 3).

#### S1.4 Documented response, scoring rule, and missing data

The documented response was the baseline minus the first post-index pain value, with meaningful relief at a reduction of at least 2 points (primary) or at least 30%. Response analyses required an interpretable baseline of 1 to 10 (eTable 8 reports the excluded counts and their reassessment rate). Because more than one post-treatment score could exist, the relief rate was recomputed using the last and the lowest (best) post-index value (eTable 6). The population relief rate was estimated by complete-case analysis, by inverse-probability-of-reassessment weighting (IPW) under a missing-at-random assumption (weights truncated at the 99th percentile; diagnostics in eTable 7), and by missing-not-at-random (MNAR) bounding that swept the assumed relief rate among never-reassessed patients across its full range, complemented by model-based imputation under MAR and under fixed MNAR offsets.

#### S1.5 Clustering and software

Because patients could contribute multiple visits, all key estimates were repeated using one visit per patient and with a patient-clustered bootstrap. Analyses used Python 3.9 with pandas 2.3, scikit-learn 1.6, SciPy 1.13, statsmodels 0.14, and matplotlib 3.9; random seeds were fixed (20260602 and 42).

#### S1.6 External validation in MC-MED

The three core analyses (reassessment completeness, the score-selection relief estimate, and the at-treatment predictability of whether a reassessment occurs) were repeated unchanged in MC-MED, a de-identified ED database from a separate US academic medical center on a different EHR. Headache visits were identified by triage chief complaint, the treated subcohort received a qualifying analgesic, and serial pain values were taken from the MC-MED numerics stream. The index treatment time was the medication administration time (First_admin_time) rather than the dispense time used in MIMIC-IV-ED; MC-MED date-shifting preserves within-visit intervals, so treatment-to-reassessment timing was valid. The at-treatment selection model used baseline pain, age, sex, arrival mode, and triage acuity, with the same 5-fold cross-validated out-of-fold AUROC as the primary analysis. Results are summarized in eTable 11.

### S2 Supplementary Tables

eTable 1. Case-identification codes and terms.

| Criterion | Codes or terms |
|---|---|
| Chief complaint (regex) | headache; head ache; migraine; cephalgia; cephalalgia |
| ICD-9 diagnosis | 346.x (migraine); 307.81 (tension-type); 339.x (other headache syndromes); 784.0 (headache) |
| ICD-10 diagnosis | G43 (migraine); G44 (other headache disorders); R51 (headache) |

eTable 2. Qualifying analgesic and abortive medication classes. Opioids were ordered first in classification so combination products containing an opioid were classified as opioid.

| Class | Matching terms (medication name) |
| --- | --- |
| Opioid | morphine, hydromorphone, oxycodone, hydrocodone, fentanyl, codeine, tramadol, oxymorphone, meperidine, butorphanol, nalbuphine, tapentadol |
| Dopamine antagonist | metoclopramide, prochlorperazine, chlorpromazine, droperidol, promethazine |
| NSAID or ketorolac | ketorolac, ibuprofen, naproxen, ketoprofen, diclofenac, indomethacin, aspirin |
| Triptan | sumatriptan, rizatriptan, zolmitriptan, eletriptan, naratriptan, frovatriptan, almotriptan |
| Dihydroergotamine | dihydroergotamine, ergotamine |
| Magnesium | magnesium |
| Acetaminophen | acetaminophen, paracetamol |

eTable 3. Adjusted odds ratios for a documented post-treatment reassessment, full model (n = 13,682). Per unit for continuous factors, versus the reference for indicators; P values Benjamini-Hochberg adjusted. Reference categories: race and ethnicity, Asian; arrival shift, day; first agent, acetaminophen.

| Factor | OR | 95% CI | Adjusted P |
| --- | --- | --- | --- |
| ED length of stay (per hour) | 1.11 | [1.10, 1.13] | <0.001 |
| No. of analgesic classes (per class) | 1.29 | [1.21, 1.38] | <0.001 |
| Triage acuity (per less-acute level) | 0.79 | [0.74, 0.85] | <0.001 |
| Night arrival | 1.40 | [1.22, 1.60] | <0.001 |
| Evening arrival | 1.17 | [1.07, 1.28] | 0.002 |
| First agent: dopamine antagonist | 1.20 | [1.07, 1.35] | 0.007 |

| Factor | OR | 95% CI | Adjusted P |
|---|---|---|---|
| Female sex | 1.10 | [1.01, 1.21] | 0.099 |
| Any parenteral agent | 0.88 | [0.77, 0.99] | 0.099 |
| Baseline pain (per point) | 1.01 | [1.00, 1.03] | 0.261 |
| First agent: opioid | 1.12 | [0.92, 1.37] | 0.483 |
| Hispanic ethnicity | 1.11 | [0.88, 1.42] | 0.662 |
| ED census at treatment (per patient) | 0.99 | [0.98, 1.01] | 0.834 |
| Age (per year) | 1.00 | [1.00, 1.00] | 0.834 |
| Black race | 1.02 | [0.82, 1.28] | 0.943 |
| First agent: NSAID | 1.01 | [0.91, 1.13] | 0.943 |
| Ambulance arrival | 0.99 | [0.89, 1.09] | 0.943 |
| Any opioid given | 1.00 | [0.84, 1.18] | 0.980 |

eTable 4. Reassessment completeness by time window and by clinical stratum (treated subcohort). Percentages with 95% Wilson CIs; the at-treatment and full-model AUROCs are 0.566 (95% CI 0.555-0.578) and 0.636 (0.626-0.647).

| Stratum | n | Any time, % (95% CI) | Within 2 h, % (95% CI) |
|---|---|---|---|
| All treated | 13,682 | 77.1 (76.4-77.8) | 47.9 (47.1-48.8) |
| Within 30 min | - | 17.3 (16.7-17.9) | - |
| Within 60 min | - | 27.5 (26.8-28.3) | - |
| Within 90 min | - | 38.5 (37.7-39.3) | - |
| Within 180 min | - | 60.7 (59.9-61.5) | - |
| Migraine-coded | 1,405 | 76.9 (74.7-79.1) | 48.0 (45.4-50.6) |
| Nonspecific headache | 12,277 | 77.1 (76.4-77.9) | 47.9 (47.1-48.8) |
| Headache-directed therapy | 9,137 | 78.5 (77.7-79.4) | 47.6 (46.6-48.6) |

| Stratum | n | Any time, % (95% CI) | Within 2 h, % (95% CI) |
|---|---|---|---|
| Chief complaint only | 2,811 | 79.0 (77.5-80.5) | 50.1 (48.2-51.9) |
| Diagnosis-coded | 10,871 | 76.6 (75.8-77.4) | 47.4 (46.5-48.3) |
| Discharged home | 10,597 | 76.2 (75.4-77.0) | 47.7 (46.8-48.7) |

eTable 5. Comparator painful presentations, reassessment completeness among treated visits. Headache is comparable to other common painful presentations and below protocol-monitored chest pain.

| Presentation | n treated | Any time, % (95% CI) | Within 2 h, % (95% CI) |
|---|---|---|---|
| Headache | 13,682 | 77.1 (76.4-77.8) | 47.9 (47.1-48.8) |
| Back pain | 13,126 | 75.6 (74.9-76.3) | 47.1 (46.3-48.0) |
| Abdominal pain | 31,243 | 80.6 (80.2-81.1) | 46.8 (46.2-47.3) |
| Renal colic | 5,033 | 79.2 (78.0-80.3) | 43.6 (42.2-45.0) |
| Chest pain | 17,309 | 86.3 (85.8-86.8) | 60.4 (59.6-61.1) |

eTable 6. Documented meaningful (at least 2-point) relief under each missingness assumption and scoring rule (n = 11,987 treated visits with interpretable baseline pain; 2,706 unmeasured). The bounding rows span the full range of unobservable outcomes.

| Estimator, assumption, or scoring rule | Relief (%) |
|---|---|
| Complete-case, first post-treatment score | 66.9 |
| Complete-case, last post-treatment score | 81.0 |
| Complete-case, best (lowest) post-treatment score | 83.4 |
| Inverse-probability-weighted (missing at random) | 67.5 |
| Model-based imputation (missing at random) | 67.5 |

| Estimator, assumption, or scoring rule | Relief (%) |
|---|---|
| Imputation, relief lowered 20 pp in unmeasured | 63.0 |
| Imputation, relief lowered 40 pp in unmeasured | 58.5 |
| Bound: none of the unmeasured improved | 51.8 |
| Bound: half of the unmeasured improved | 63.1 |
| Bound: all of the unmeasured improved | 74.4 |
| Migraine-coded, complete-case (first) | 74.1 |
| Nonspecific headache, complete-case (first) | 66.1 |
| Headache-directed therapy, complete-case (first) | 68.5 |

eTable 7. Inverse-probability-weighting diagnostics (reassessed visits with interpretable baseline pain). Weights are well behaved and covariate balance is achieved after weighting.

| Diagnostic | Value |
|---|---|
| Predicted reassessment probability, min to max | 0.45 to 1.00 |
| Weight, median / 99th percentile / max | 1.29 / 1.71 / 2.02 |
| Effective sample size (of 9,281 reassessed) | 9,162 |
| Calibration slope (intercept) | 0.96 (0.03) |
| Standardized mean difference, baseline pain (unweighted / weighted) | 0.017 / -0.001 |
| Standardized mean difference, ED length of stay (unweighted / weighted) | 0.083 / 0.001 |
| Standardized mean difference, No. of classes (unweighted / weighted) | 0.056 / -0.003 |

eTable 8. Reassessed versus non-reassessed visits and missing-baseline accounting (treated subcohort). Categorical factors use the chi-square test with Cramer V; continuous factors use the Kruskal-Wallis test with epsilon-squared.

| Factor or count | Reassessed | Not reassessed | P | Effect size |
|---|---|---|---|---|
| ED length of stay (median, h) | 6.2 | 5.2 | <0.001 | epsilon-squared 0.027 |
| No. of analgesic classes (median) | 2 | 1 | <0.001 | epsilon-squared 0.011 |
| Baseline pain (median) | 7 | 7 | 0.002 | epsilon-squared 0.001 |
| Any opioid given | 81.1% | 75.7% | <0.001 | Cramer V 0.057 |
| Any parenteral agent | 77.3% | 76.2% | 0.279 | Cramer V 0.009 |
| Treated visits, total | 13,682 | | | |
| Excluded: baseline missing / zero / >10 | 254 / 1,305 / 136 | | | |
| Reassessment rate, included vs excluded | 77.4% | 75.0% | | |

eTable 9. Reporting-guideline coverage. The study followed STROBE (cross-sectional) with the RECORD extension and TRIPOD for the prediction component.

| Guideline item area | Location |
|---|---|
| Design, setting, dates | Methods, Study Design and Setting |
| Eligibility and case definition | Methods, Cohort and Comparators; eTable 1 |
| Variables and data sources | Methods, Treatment and Pain Reassessment; eTable 2 |
| Statistical methods, missing data | Methods, Statistical Analysis; S1.3 to S1.5 |
| Model specification and validation (TRIPOD) | S1.3; eTable 3; eTable 4 footnote |
| Participants and descriptive data | Results, Cohort; Table 1 |
| Outcome data and estimates | Results; eTables 4 to 8 |
| Limitations and generalizability | Discussion, Limitations |

eTable 10. Post-treatment score counts and the mechanism of best-score inflation (reassessed visits with interpretable baseline pain). The best-score rule equals the first-score rule when only one score exists and inflates relief only as the number of post-treatment scores grows, while first-score relief

falls, indicating that additional measurement is prompted by slower improvement.

| Group | n | Relief, first score (%) | Relief, best score (%) | Inflation (pp) |
|---|---|---|---|---|
| 1 post-treatment score | 4,973 | 79.1 | 79.1 | 0.0 |
| 2 post-treatment scores | 2,348 | 58.0 | 86.2 | 28.2 |
| 3 or more post-treatment scores | 1,960 | 46.7 | 90.6 | 43.9 |

Among reassessed visits the median number of post-treatment scores was 1 (IQR, 1 to 2); 53.3% had one, 46.7% had two or more, and 21.3% had three or more. The last documented score fell a median of 176 minutes (IQR, 95 to 321) after treatment.

eTable 11. External validation in MC-MED versus the MIMIC-IV-ED primary cohort. The score-selection sensitivity and the near-chance at-treatment predictability replicated; reassessment completeness was poorer in MC-MED and its absolute magnitude did not port across systems.

| Measure | MC-MED | MIMIC-IV-ED |
|---|---|---|
| Chief-complaint headache visits, No. | 4,843 | 19,501 |
| Treated with a qualifying analgesic, No. | 2,257 | 13,682 |
| Reassessed responders for relief analysis, No. | 211 | 9,281 |
| Meaningful relief, first post-treatment score, % | 71.1 | 66.9 |
| Meaningful relief, last post-treatment score, % | 80.6 | 81.0 |
| Meaningful relief, best (lowest) score, % | 83.4 | 83.4 |
| At-treatment AUROC for a documented reassessment | 0.501 | 0.566 |
| Completeness, any time before disposition, % (95% CI) | 11.8 (10.6 to 13.2) | 77.1 (76.4 to 77.8) |
| Completeness, within 120 min, % | 8.8 | 47.9 |
| Completeness, within 60 min, % | 6.0 | 27.5 |

MC-MED uses medication administration time (First_admin_time); MIMIC-IV-ED uses dispense time. The cohort is chief-complaint headache in both. Date-shifting in MC-MED preserves within-visit intervals.

### S3 Supplementary Figures

eFigure 1. Distribution of the triage pain score in the headache cohort. Bars show the percentage of visits at each integer of the 0 to 10 numeric rating scale; the heaped values at 0, 5, 8, and 10 are shaded darker, illustrating the coarse, non-continuous behavior of the instrument that anchors the documented response.

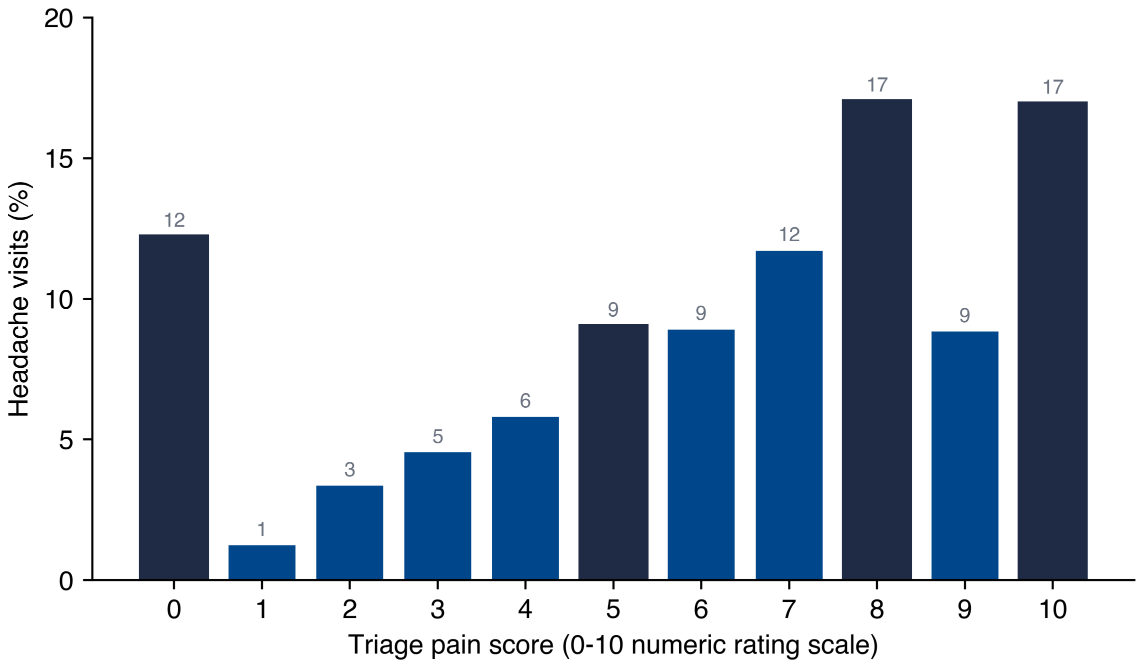


eFigure 2. Adjusted odds ratios for a documented post-treatment reassessment (full model, n = 13,682). Points are adjusted odds ratios with 95% confidence intervals; the dashed line marks the null. Baseline pain (highlighted) shows no association. Areas under the curve are shown for the full model and for the at-treatment model that excludes post-index visit course. Reference categories: race and ethnicity, Asian; arrival shift, day; first agent, acetaminophen.

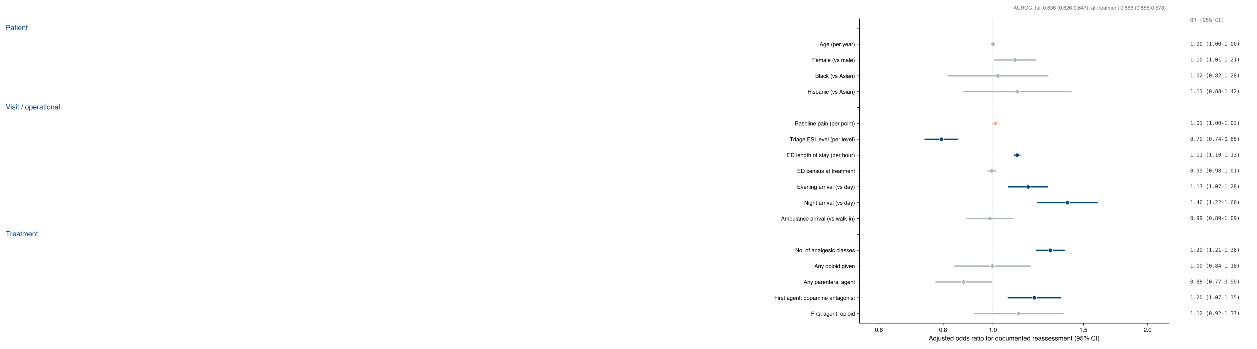


eFigure 3. Identification range of the population relief rate under missing-not-at-random assumptions. The line shows the population proportion achieving a reduction of at least 2 points as a function of the assumed relief rate among never-reassessed patients (an assumption, not an estimate); the shaded region is the identified band. The complete-case (66.9%) and inverse-probability-weighted (67.5%) estimates are marked. The band spans 51.8% to 74.4%, a width equal to the unmeasured fraction.

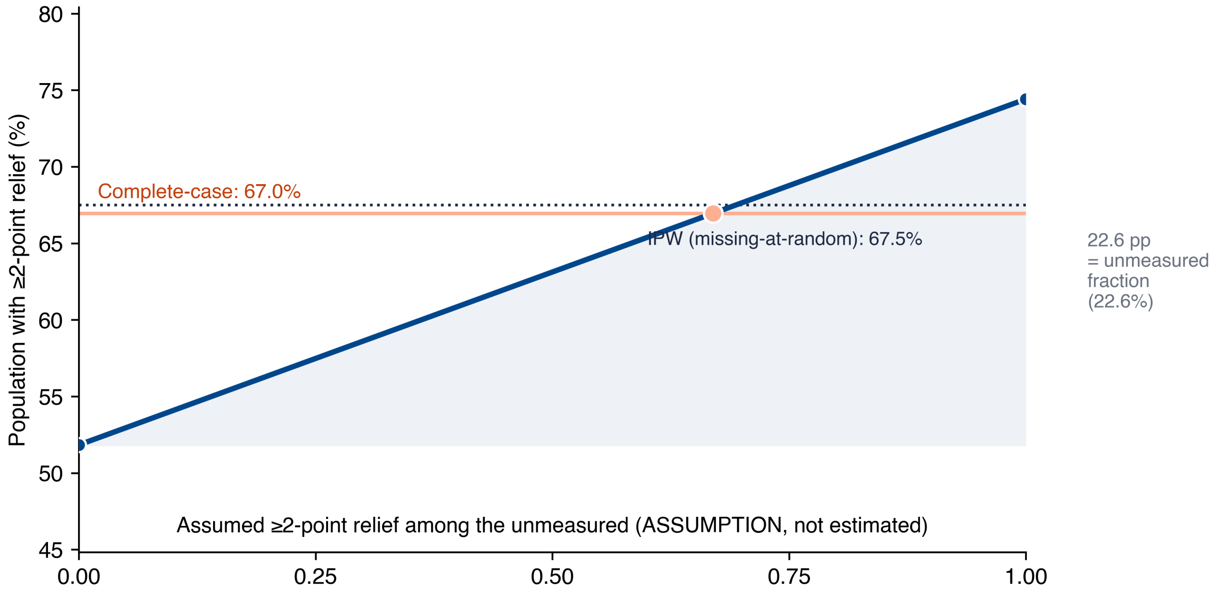


### S4 Endpoint Codebook

- **Treated visit.** At least one dispense of a qualifying analgesic or abortive agent (eTable 2).

- **Index treatment time.** Earliest qualifying dispense time within the visit.
- **Baseline pain.** Last numeric 0 to 10 pain value at or before the index time (serial value preferred, triage value otherwise).
- **Documented reassessment.** At least one numeric pain value recorded after the index time within the same ED stay; window variants require it within 30 to 180 minutes.
- **Documented response.** Baseline pain minus a post-index pain value (first, last, or best).
- **Meaningful relief.** Documented response of at least 2 points (primary) or at least 30% (secondary).
- **Response cohort.** Treated visits with an interpretable baseline pain of 1 to 10.

### S5 Code and Data Availability

The analysis used the publicly available, de-identified MIMIC-IV-ED version 2.2 database, accessible to credentialed users through PhysioNet under its data use agreement. The authors did not have special access privileges. Analysis code (cohort construction, measurement, modeling, bounding, comparator analysis, and figures) is available at https://github.com/Alon-Gorenshtein/study_headache_unmeasured_relief. The derived analytic dataset cannot be redistributed under the data use agreement; it is reproducible from the source tables with the provided code. Correspondence: Alon Gorenshtein, MD.
